# Health system transitions and engagement for patients with opioid use disorder

**DOI:** 10.64898/2026.09.22.26363750

**Authors:** Meredith Niess, Kendrick Kennedy, Victoria Glunt, Joshua Burrows, Ben Goldstein, Amanda Brucker, Carl Mhina, Catherine Staton, Michael Pignone

**Affiliations:** Department of Medicine, Duke University Medical Center; Department of Emergency Medicine, Duke University Medical Center; Collaborative to Advance Clinical Health Equity, Duke Health and Duke University School of Medicine; Department of Biostatistics & Bioinformatics, Duke University School of Medicine; Department of Population Health Sciences, Duke University, Durham, North Carolina, USA

## Abstract

**Background:** Opioid Use Disorder (OUD) imposes a rising mortality and health system burden, with opioid-related deaths increasing from 8,400 in 2000 to 110,037 in 2023. Medications for opioid use disorder (MOUD) reduce mortality and morbidity, yet only 25.1% of people with OUD are treated with these medications, and inpatient MOUD initiation is inconsistent. Inpatient and Emergency Department (ED) treatment initiation and connection to outpatient care is suboptimal, missing critical opportunities to engage at-risk populations during acute care visits. Our goals were to determine the proportion of patients with OUD treated with MOUD during a hospital/ED stay, and the proportion successfully connected to outpatient care.

**Methods:** This retrospective cohort study included patients ≥18 years with an OUD diagnosis at Duke University Hospital and Duke Regional Hospital (7/1/2023–12/31/2025) who had an inpatient or ED encounter with an associated OUD diagnosis code. Primary outcomes were buprenorphine at discharge, 30-day acute care return, and 30-day outpatient follow-up. Adjusted modified Poisson regression models with cluster-robust standard errors evaluated associations with age, sex, race/ethnicity, insurance, area deprivation index, encounter type, and comorbidity score.

**Results:** Of 5,390 patients with OUD, 42.1% (2,267) had an acute care encounter, generating 3,893 encounters. Neither buprenorphine nor methadone was prescribed at discharge for 46.0% of encounters. Acute care return occurred in 33.6% and outpatient follow-up in 32.2% of encounters within 30 days; only 7.4% of follow-ups included a buprenorphine prescription. ED encounters had lower discharge prescribing than inpatient encounters (RR 0.72, 95% CI 0.61–0.86). Black patients had higher 30-day acute care return than White patients (RR 1.16, 95% CI 1.02–1.30)

**Conclusions:** Acute care encounters represent repeated, missed opportunities for MOUD initiation. Disparities were concentrated in post-discharge return and follow-up rather than at initial prescribing, and attrition occurred at each transition along the screening-treatment-referral pathway.

## Introduction

Opioid Use Disorder (OUD) is the third leading substance use disorder worldwide ^1^. National Centers for Disease Control and Prevention (CDC) data show an exponential rise in opioid-related deaths, from 8,400 in 2000 to 110,037 in 2023, with mortality rates rising from 1.2 to 5.9 per 100,000 over a comparable period (1998–2016)^3^. North Carolina has remained above the national average in opioid overdose deaths, with an estimated rate of 21.7 per 100,000 in 2025^4^, compared to the U.S. national rate of 20.5 per 100,000^5^. Independent of mortality, the prevalence of OUD places an overwhelming burden on health systems. Hospitalization rates for OUD rose from 59.8 to 190.7 per 100,000 between 1998 and 2016^3^, and OUD is estimated to impose an annual health system burden exceeding $89–$107 billion in direct medical expenditures and hospital costs in the US ^6^.This burden is compounded by longer hospital stays patients with OUD have a length of stay ranging from 19%–23% longer than patients hospitalized without OUD ^7^, and they experience the highest unplanned 30-day readmission rates of any patient population, at nearly 40% ^8^. This strain is especially evident in Emergency Departments (ED). Out of 234 million adult ED visits in the United States between 2016 and 2017, more than 2.88 million were opioid-related^9^.

Medication management of opioid use disorder (MOUD) with Food and Drug Administration (FDA) approved medications including methadone, buprenorphine, and naltrexone reduces mortality and morbidity^1^. Furthermore, buprenorphine and methadone significantly reduce overdose-related and overall mortality^10^. Yet, as of 2022, only 25.1% of people in the US with OUD have been treated with these medications. Hospital-based cohort studies have also shown that inpatient MOUD care initiations are low, ranging from as low as 13% to 45 ^10,11^. While buprenorphine and naloxone are prescribed in office-based settings and can be taken at home, outpatients with OUD are only able to receive methadone at federally regulated clinics (OTPs), creating further barriers to successful MOUD maintenance. Beyond these low rates of MOUD care, significant disparities exist in who receives treatment. Nationally, only 25% of adults who need OUD treatment in 2022 received MOUD, with the rates being lower among Black and Hispanic adults and among women than men ^12^. This gap is wider in specific populations. Among Medicare disability beneficiaries, Black patients received buprenorphine far less often than White patients (9.1% vs 21.6% in 2016; 14.1% vs 25.5% in 2019), despite comparable outpatient visit frequency ^13^. These disparities exist even when screening rates are high. In a cohort study of 12 million adults across 1,300 clinics, Spanish preferring Latino patients had the highest rates of screening, despite this, all minoritized groups had lower odds of being prescribed addiction treatment medications than non-Hispanic White patients (Chan et al. 2026).

Nationally, inpatient and Emergency Department initiation of treatment and connection to outpatient care is suboptimal, missing critical opportunities to engage our most at-risk populations during acute care visits^17–24^. Therefore, our project’s overall goals were 1) to determine the proportion of patients in our health system with OUD treated with MOUD during a hospital or ED stay, and 2) to determine the proportion of OUD patients successfully connected to outpatient care.

## Methods

### Study Design and Setting

This is a retrospective, cohort study of patients seen at Duke University Hospital and Duke Regional Hospital in the Duke University Medical Center system. Duke University Hospital (DUH) has 1,062 inpatient beds and offers regular and intensive care inpatient units as well as comprehensive diagnostic and therapeutic facilities, including a regional emergency/trauma center. Duke Regional Hospital (DRH) is a 388-bed acute care community hospital serving residents of Durham and surrounding counties.

### Populations

The study cohort included all patients in the Duke Health system with a billing encounter or problem list OUD diagnosis between 7/1/2023 and 12/31/2025, and who were greater than 18 years of age as of 7/1/2023. We then created an acute care population for patients who an inpatient or Emergency Department encounter at either hospital during the study period with an OUD diagnosis code associated with the encounter (F11).

### Outcomes

All variables were extracted from the electronic medical record. The primary outcomes were buprenorphine at discharge, 30-day acute care return visit, and 30-day follow-up outpatient treatment. A patient was considered to have a buprenorphine at discharge if either buprenorphine or buprenorphine-naloxone was ordered for outpatient fill or on the patient’s medication reconciliation list at discharge. Although naltrexone is an FDA approved treatment for OUD, it is not commonly used in practice for OUD due to increased overdose risk with treatment cessation and high rates of use or Alcohol Use Disorder. Methadone orders were not included in the primary outcome due to the federal regulatory restrictions on methadone. A 30-day acute care return visit was defined as any inpatient or ED visit within 30 days of the index visit. A 30-day follow-up outpatient treatment visit was defined as an outpatient visit within 30 days of the index visit with a buprenorphine or buprenorphine-naloxone order for outpatient fill in one of the following settings: Duke Primary Care, Duke Outpatient Clinic, Duke Family Medicine, Infectious Diseases, Medicine Pediatrics, Obstetrics and Gynecology, or Psychiatry.

### Covariates

We adjusted for the following potential confounders: age at first encounter, sex, race/ethnicity, insurance status at encounters during the study period (public, private or self-pay), Area of Deprivation Index (ADI) ^25,26^, (1-5 or 6-10), encounter type (inpatient or ED) and the comorbidity score (0-2, 3-4 or 5-9).

### Statistical Analysis

Patient characteristics and outcomes were summarized descriptively. Continuous variables were summarized using means, medians, standard deviations, quartiles, and ranges. Categorical variables were summarized using frequencies and percentages. Absolute standardized mean differences (ASMDs) are reported to quantify imbalances in patient characteristics. Adjusted and unadjusted modified Poisson regression models with cluster-robust standard errors were used to evaluate the association between buprenorphine at discharges and the following comparators: age (<30 vs. ≥30), sex, race/ethnicity, insurance type, area deprivation index (ADI) (1-5 vs. 6-10), and comorbidity score (0-2, 3-4, 5-9). A similar analysis was performed for the 30-day acute care return visit outcome. The analysis for 30-day return visit also evaluated the association with index encounter buprenorphine at discharge (Yes/No). The adjusted models included main effects for the comparator group variable of interest as well as all other comparator group variables. Adjusted and unadjusted relative risks are reported with Wald 95% confidence intervals (CIs). Analysis of the primary outcomes was performed at the encounter level. No adjustment was made for multiple comparisons. All statistical analysis was performed in R 4.4.1 (R Core Team 2024).

## Results

### Characteristics of Study Participants

A total of 5,390 patients with OUD diagnoses were identified as receiving care within the Duke Health system from July 2023 to December 2025. Of the 5,390 patients with an OUD diagnosis, 42.1% (2,267/5,390) had an inpatient or emergency department visit during the 30-month study period, forming our OUD/Acute population (**Figure 1**). Most patients had one (69.7%) or two (15.9%) acute encounters over the 30-month period, with only 123 (5.4%) having 5 or more encounters **(Appendix Table1)**.

**Figure 1:**
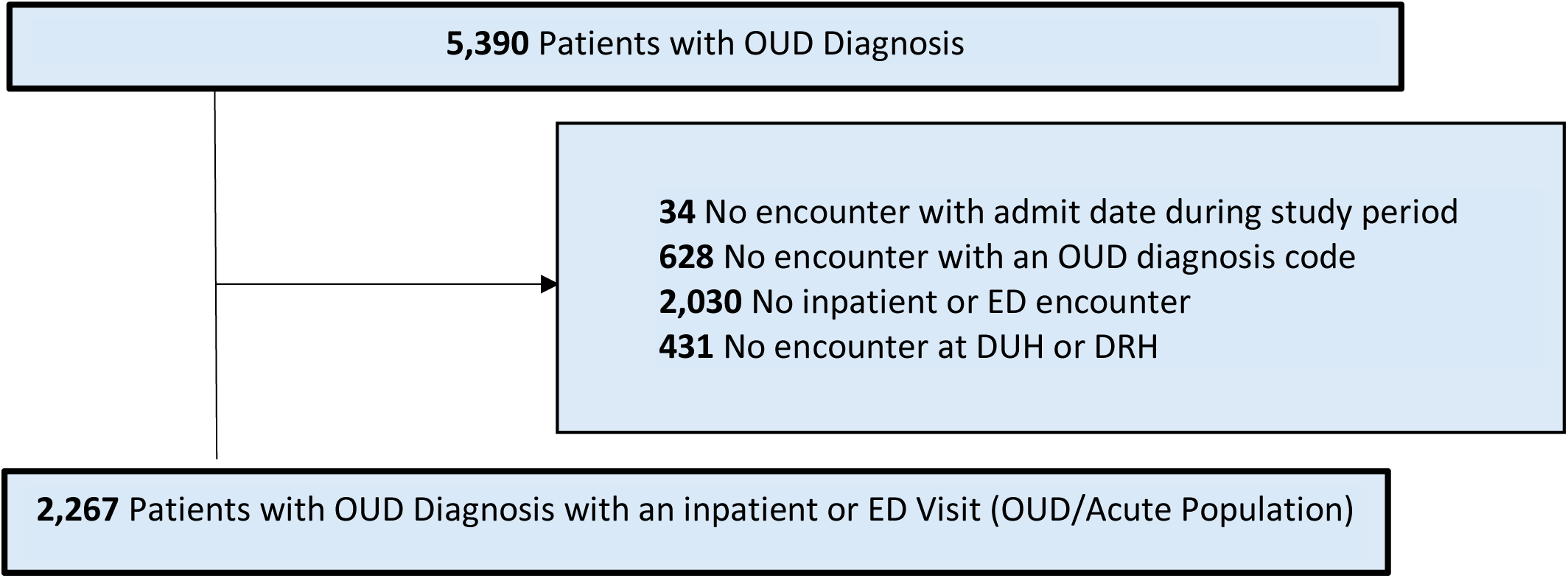
Study Flow Diagram after identifying patients with acute care encounters

Patients with an acute encounter were younger (mean age 45.4 vs. 51.5 years) and the encounter were more often for males (55.7% vs. 48.8%) than those without acute encounters. Patients with acute encounters were also more likely to be Black (34.9% vs. 27.0%) and less likely to be White (59.4% vs 68.5 %) (**Table 1**). Insurance status also differed between the groups. Patients with an acute encounter were more likely to have Medicaid (45.2% vs 25.7%) or self-pay status (10.9% vs 7.7% and less likely to have Medicare (24.7% vs 41.7%) or commercial.

**Table 1:** Sociodemographic and clinical characteristics of patients enrolled in the study.

|  | Overall<br>N = 5,390 | Patients with an<br>acute encounter<br>N = 2,267 | Patients without<br>an acute encounter<br>N = 3,123 | ASMD |
| --- | --- | --- | --- | --- |
| Age at first encounter |  |  |  |  |
| Mean (SD), y | 48.9 (15.5) | 45.4 (14.4) | 51.5 (15.8) | 0.40 |
| Sex |  |  |  | 0.14 |
| Female | 2,604 (48.3%) | 1,005 (44.3%) | 1,599 (51.2%) |  |
| Male | 2,786 (51.7%) | 1,262 (55.7%) | 1,524 (48.8%) |  |
| Race |  |  |  | 0.19 |
| American Indian or Alaska Native | 48 (0.9%) | 22 (1.0%) | 26 (0.8%) |  |
| Asian | 18 (0.3%) | 10 (0.4%) | 8 (0.3%) |  |
| Black | 1,634 (30.3%) | 792 (34.9%) | 842 (27.0%) |  |
| Native Hawaiian or Other Pacific Islander | 3 (0.1%) | 1 (0.0%) | 2 (0.1%) |  |
| White | 3,486 (64.7%) | 1,347 (59.4%) | 2,139 (68.5%) |  |
| Other | 70 (1.3%) | 40 (1.8%) | 30 (1.0%) |  |
| Unknown | 131 (2.4%) | 55 (2.4%) | 76 (2.4%) |  |
| Hispanic ethnicity |  |  |  | 0.09 |
| Yes | 148 (2.7%) | 74 (3.3%) | 74 (2.4%) |  |
| No | 5,029 (93.3%) | 2,126 (93.8%) | 2,903 (93.0%) |  |
| Unknown | 213 (4.0%) | 67 (3.0%) | 146 (4.7%) |  |
| Patient preferred language |  |  |  | 0.03 |
| English | 5,354 (99.3%) | 2,253 (99.4%) | 3,101 (99.3%) |  |
| Spanish | 24 (0.4%) | 11 (0.5%) | 13 (0.4%) |  |
| Other | 12 (0.2%) | 3 (0.1%) | 9 (0.3%) |  |
| Insurance |  |  |  | 0.42 |
| Medicare | 1,861 (34.5%) | 560 (24.7%) | 1,301 (41.7%) |  |
| Medicaid | 1,829 (33.9%) | 1,025 (45.2%) | 804 (25.7%) |  |
| Other public | 27 (0.5%) | 16 (0.7%) | 11 (0.4%) |  |
| Commercial | 991 (18.4%) | 328 (14.5%) | 663 (21.2%) |  |
| Self-pay | 486 (9.0%) | 247 (10.9%) | 239 (7.7%) |  |
| Other | 144 (2.7%) | 91 (4.0%) | 53 (1.7%) |  |
| Missing | 52 (1.0%) | 0 (0.0%) | 52 (1.7%) |  |
| County of residence |  |  |  | 0.53 |
| Durham | 1,776 (32.9%) | 1,067 (47.1%) | 709 (22.7%) |  |
| Adjacent to Durham (Granville, Wake, Chatham, Orange, Person) | 1,735 (32.2%) | 568 (25.1%) | 1,167 (37.4%) |  |
| Other NC county | 1,515 (28.1%) | 504 (22.2%) | 1,011 (32.4%) |  |
| Out of state | 171 (3.2%) | 79 (3.5%) | 92 (2.9%) |  |
| Missing | 193 (3.6%) | 49 (2.2%) | 144 (4.6%) |  |
| Area Deprivation Index (ADI) |  |  |  | 0.20 |
| 1-5 | 2,899 (53.8%) | 1,091 (48.1%) | 1,808 (57.9%) |  |
| 6-10 | 1,998 (37.1%) | 963 (42.5%) | 1,035 (33.1%) |  |
| Missing | 493 (9.1%) | 213 (9.4%) | 280 (9.0%) |  |

### Acute Care Population

A total of 2,267 (42.1) of patients with an OUD diagnosis had an acute care encounter. Most patients had at least one acute OUD inpatient encounter (1,837/2,267; 81.0%) (**Appendix Table 2**). A minority of patients had an emergency department acute OUD encounter only (430/2,267; 19.0%).

**Table 2:** Encounter-level outcomes for patients with an acute encounter.

| Acute DUH/DRH OUD encounters for patients with an acute OUD encounter<br>N = 3,893 |  |
| --- | --- |
| **MOUD prescription at discharge (Outpatient fill or on medication reconciliation list)** |  |
| MOUD prescription | 3893 |
| Buprenorphine or buprenorphine-naloxone | 1,322 (34.0%) |
| Methadone | 755 (19.4%) |
| Both | 26 (0.7%) |
| Neither (opportunity) | 1,790 (46.0%) |
| **Follow-up encounters within 30 days of hospital discharge** |  |
| Inpatient or ED (i.e. Acute care return visit) | 1,307 (33.6%) |
| Outpatient | 1,252 (32.2%) |
| **Follow-up encounters within 30 days of hospital discharge with buprenorphine or buprenorphine-naloxone at follow-up** |  |
| Outpatient | 287 (7.4%) |
| **Follow-up encounters within 30 days of discharge with OUD diagnosis code** |  |
| Inpatient or ED | 475 (12.2%) |
| Outpatient | 149 (3.8%) |

#### Sociodemographic Characteristics

The mean patient age was 46.7 (SD 14.5) and 39.8 (SD 12.9) among patients who had an OUD inpatient encounter and patients who did not, respectively (**Appendix Table 2**). The rate of self-pay insurance was 7.2% and 26.5% among patients who had an OUD inpatient encounter and patients who did not, respectively. The comorbidity rates were higher among patients who had an OUD inpatient encounter than patients who did not.

#### Clinical diagnosis and comorbidities

The most common comorbidities for the OUD population with an acute visit were pain (78.1%), psychiatric conditions (62.9%), complications associated with injected drug use (60.1%), and renal disease (36.6%), followed by liver disease (19.3%), Hepatitis C (18.6%), opioid overdose (13.4%), and HIV infection (2.2%). Comorbidity scores varied across the cohort with 18% having 5-9 comorbidities, and 42.4% having 3-4 comorbidities, with 39.2%having only 0-2 comorbidities (**Appendix Table 2**).

#### Clinical diagnosis and comorbidities

The most common comorbidities for the OUD population with an acute visit were pain (78.1%), psychiatric conditions (62.9%), complications associated with injected drug use (60.1%), and renal disease (36.6%), followed by liver disease (19.3%), Hepatitis C (18.6%), opioid overdose (13.4%), and HIV infection (2.2%). Comorbidity scores varied across the cohort with 18% having 5-9 comorbidities, and 42.4% having 3-4 comorbidities, with 39.2%having only 0-2 comorbidities (**Appendix Table 2**.

#### Acute Care Encounters and Outcomes

Among the 2,267 patients in the OUD/Acute population, there were 3,893 acute OUD encounters **(Table 2).** Of these 3893 visits, for 46.0 of these encounters, patients did not receive a buprenorphine or buprenorphine-naloxone, nor a methadone discharge plan compared to % who did. 33.6% (n=1307) of encounters had an acute care return visit (i.e., an inpatient or ED visit), and 32% (n=1252) of encounters had an outpatient follow up within 30 days of discharge Of those with an outpatient follow-up in the Duke ambulatory setting within 30 days, only 7.4% (n=287) received a prescription at this follow-up visit for initiation or continuation of buprenorphine or buprenorphine-naloxone, representing less than 22% of those discharged with the medication, and less than 10% ( )of those discharged with either buprenorphine or buprenorphine-naloxone or no MOUD.

### Unadjusted and Adjusted Model Results

#### Buprenorphine at discharge

Comparing patients age ≥30 to patients age <30, the adjusted estimated relative rate of buprenorphine at discharge was 1.31 (95% CI: 1.05, 1.64). The unadjusted estimate was similar. Patients with an emergency department encounter had a lower relative rate compared to those with an inpatients encounter (0.72, 95% CI 0.61-0.86) (**Table 3**)

**Table 3:** Model-estimated relative rates of buprenorphine at discharge across comparator groups.

|  | Unadjusted relative rate (95% CI) | Adjusted relative rate (95% CI) |
| --- | --- | --- |
| Age |  |  |
| <30 | REF | REF |
| ≥30 | 1.39 (1.11, 1.74)* | 1.31 (1.05, 1.64)* |
| Sex |  |  |
| Female | REF | REF |
| Male | 1.09 (0.94, 1.26) | 1.09 (0.94, 1.26) |
| Race/ethnicity |  |  |
| Non-Hispanic White | REF | REF |
| Hispanic | 0.82 (0.53, 1.26) | 0.90 (0.58, 1.39) |
| Non-Hispanic Black | 0.92 (0.78, 1.07) | 0.93 (0.79, 1.08) |
| Other/Unknown | 1.11 (0.86, 1.44) | 1.07 (0.84, 1.35) |
| Insurance |  |  |
| Public | REF | REF |
| Private | 1.20 (1.00, 1.44) | 1.18 (0.98, 1.42) |
| Self-pay | 1.00 (0.81, 1.23) | 1.12 (0.90, 1.38) |
| Other | 1.09 (0.83, 1.45) | 1.08 (0.80, 1.44) |
| ADI |  |  |
| 1-5 | REF | REF |
| 6-10 | 1.03 (0.89, 1.19) | 1.01 (0.88, 1.17) |
| Missing | 0.88 (0.67, 1.17) | 0.86 (0.65, 1.13) |
| Encounter type |  |  |
| Inpatient | REF | REF |
| ED | 0.71 (0.60, 0.84) | 0.72 (0.61, 0.86) |
| Comorbidity score |  |  |
| 0-2 | REF | REF |
| 3-4 | 0.97 (0.84, 1.13) | 0.92 (0.79, 1.07) |
| 5-9 | 1.12 (0.93, 1.36) | 1.04 (0.86, 1.26) |

#### Acute care return visit within 30 days

The rate of acute care return visit within 30 days among encounters with Non-Hispanic Black patients was estimated to be 1.16 (95% CI: 1.02, 1.30) times the rate among encounters with Non-Hispanic White patients **(Table 4)**. The rates of acute care return visit within 30 days among encounters with private insurance patients was 0.83 (95% CI: 0.68, 1.00) and with self-pay insurance patients was estimated to be 0.78 (0.61, 0.99) times the rate among encounters with public insurance patients. The rate of acute care return visit among ED encounters was estimated to be 1.60 (95% CI: 1.42, 1.80) times the rate among inpatient encounters. The rate of acute care return visit was also associated with the number of comorbidities; patient encounters with a 3-4 comorbidity score have 1.79 (95% CI: 1.51, 2.12), and patient encounters with a 5-9 comorbidity score have 2.34 (95% CI: 1.95, 2.81) times the rate among encounters with comorbidity score 0-2 patients.

**Table 4:**
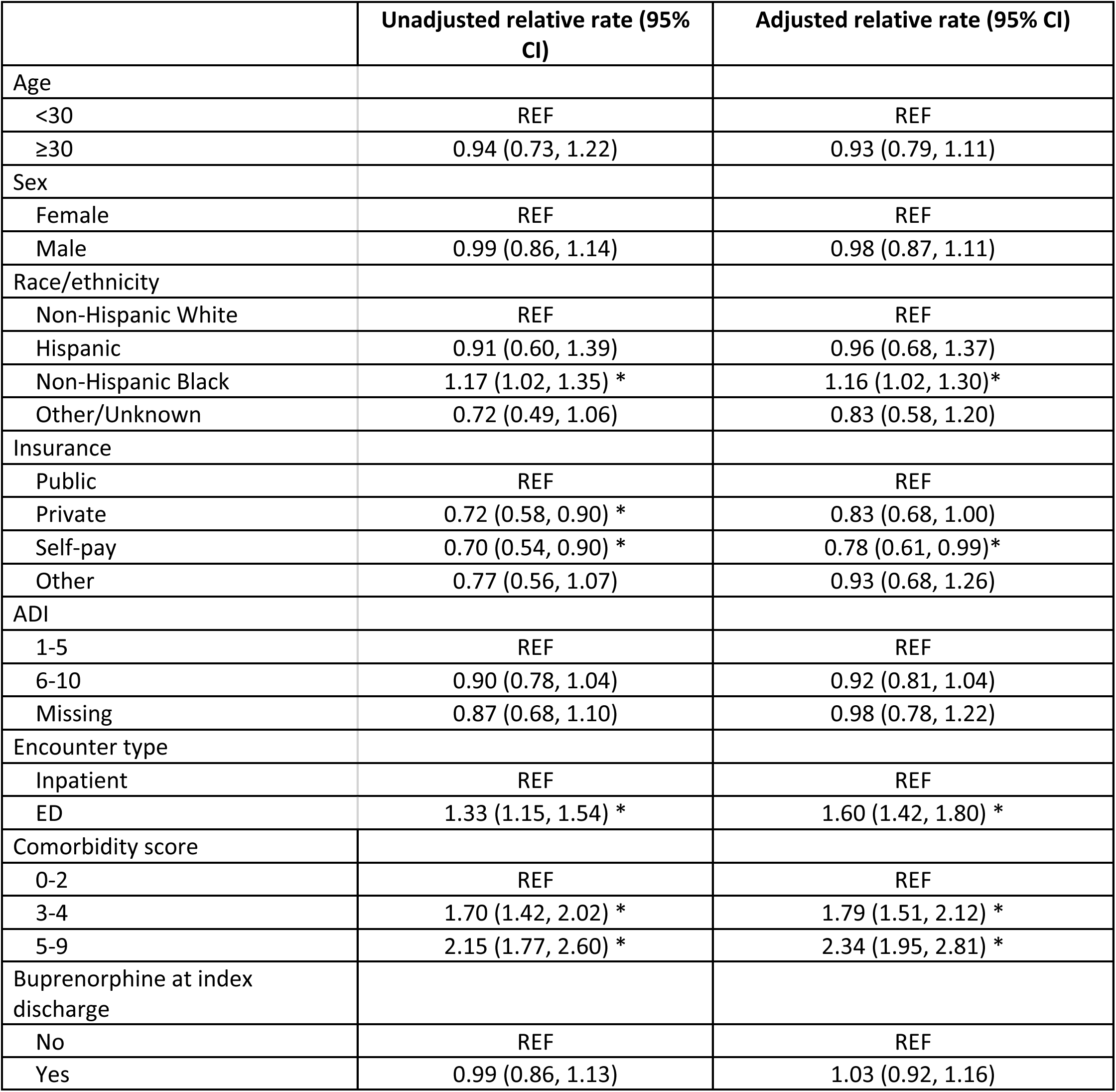
Model-estimated relative rates of acute care return visit within 30 days across comparator groups.

## Discussion

Our analysis yielded three main findings. First, patients with OUD presenting for acute care were high risk, with significant comorbidly and risk for return or “bounce back” acute care visits. This complexity and utilization pattern underscore the repeated opportunities for identification of OUD and subsequent MOUD initiation that our healthcare system has not yet fully captured. Second, disparities in this population were concentrated in acute-care return visits and 30-day follow-up rather than at the point where the decision is made to prescribe MOUD at the initial patient encounter, suggesting that inequities emerge downstream of the initial treatment decision rather than at the point of prescribing itself. Lastly, there was a marked drop-off between MOUD prescribing at initial hospital discharge and a confirmed outpatient MOUD appointment. This disparity points to a fragmented, multi-step linkage process of screening, treatment, and referral (STR). This underscores that patient attrition occurs at each transition along that pathway, rather than at a single point of failure.

Patients presenting in the acute care setting had recurrent acute care utilization, but we find that there are several missed opportunities to intervene to increase diagnosis and treatment. These missed opportunities reduce improving downstream outcomes related to return acute visits, follow-up visits for MOUD, and overall addiction severity ^27^. Of patients seen in the acute setting, a third had more than one acute encounter over the 18-month study period, and 5.4% presented five or more times. A notable pattern of recurrent, high acute contact that did not yield proper MOUD initiation and follow up. Despite this repeated contact, 47.5% of the 3,893 initial encounters in our OUD/Acute population had neither buprenorphine/buprenorphine-naloxone nor methadone recorded at discharge, underscoring a substantial, recurring missed opportunity to initiate evidence-based treatment. Addressing this gap is important because ED-initiated has been linked to fewer ED visits ^28^and greater engagement in outpatient treatment after discharge^29^. This underscores a substantial, recurring opportunity to initiate evidence-based treatment that was not consistently acted upon.

Encounter type in our study was the factor most strongly associated with receiving a discharge prescription. Patients who were discharged from the ED had a lower adjusted rate of buprenorphine or buprenorphine-naloxone prescribing than inpatient encounters. This likely reflects the ED’s shorter treatment window, the lack of a standardized workflow to support evidence-based initiation, and gaps in provider education and comfort with prescribing. Standardized workflow in our health system. have shown to be effective. Hospitalized patients in the Caring for patients of Patients with Opioid Misuse through Evidence Based Treatment (COMET) were associated with higher rates of inpatient MOUD initiation and a 23% lower rate of 30-day readmission ^30^. The takeaway here is that the ED remains an underused, and potential high-yield target for a standardized MOUD initiation protocol ^31,32^.

There was little evidence of disparity in the decision to prescribe MOUD at discharge. MOUD prescribing did not differ meaningfully by sex, race/ethnicity, or insurance status. The only meaningful differences were age and encounter type. Patients with older age and inpatients (compared to ED) were both linked to a higher likelihood of receiving MOUD. Disparities instead during acute care return within 30 days: Black patients returned more often than White patients, privately insured and self-pay patients returned less often than those who were publicly insured and returns rose with comorbidity burden and were more common after ED-only encounters. This pattern suggests that the decision to treat with MOUD is independent of patient specific factors, but what happens to a patient post-discharge is not. A plausible driver is that stable outpatient follow-up depends on factors unaccounted for at discharge like transportation access, ability to take excused time at work, and clinic capacity, among others. The higher return rate among Black patients and publicly insured patients may reflect greater reliance on the ED as a safety net access point ^33^, particularly since receiving MOUD at the initial encounter was not itself associated with a lower return rate. The lower return rate among self-pay patients likely reflects something different, this may represent cost avoidance of acute care altogether rather than better outcomes, given the direct out-of-pocket cost these patients face^34^. These patients have a direct out-of-pocket cost that may affect their desire to return to the ED. It’s worth noting that our overall prescribing and referral rates are low across the board, and low baseline rates can themselves mask disparities that may exist. These will become more visible as STR increases. Additionally, the entire OUD population in this cohort is subject to both societal and healthcare related stigma. This is further exacerbated by previous negative and traumatic healthcare experiences^35–37^. All these factors plausibly suppress prescribing, referral, and follow up rates across every subgroup examined.

One of the clearest signals in our data is the amount of attrition across the STR pathway. When looking thoroughly at the information we find that roughly a third of the initial encounters had a buprenorphine/buprenorphine-naloxone prescription at discharge. A similar share resulted in an acute care return visit within 30 days, and the same share had an outpatient follow-up visit. Unfortunately, patients who had a confirmed medication linked outpatient follow-up dropped to less than ten percent, and outpatient follow-up with an OUD-specific diagnosis code fell to one in twenty-five. Acute care visits occurred at essentially the same rate as outpatient follow up, which suggests that for a large portion of this population that the hospital/ED, not primary or specialty care, remains the default site of reengagement after an OUD related encounter. This reflects a broader pattern of attrition across the acute to outpatient pathway, with losses occurring at nearly every step between an acute encounter and patient receiving outpatient MOUD treatment. It should be noted that, not all the attrition is the same. Some of it can be addressed within the system by being thoughtful of offering treatment during acute encounters, discharging patients with a planned/scheduled outpatient follow up, understanding why patients may have no-shows to appointments. However, there are some problems that cannot be addressed by our system that include those who have received OUD treatment outside of the healthcare system, and those who are currently prescribed methadone who are required to receive treatment through a separate federally regulated clinic. This all matters as it highlights a structural hurdle that causes gaps in care. If the outpatient and acute care services are siloed, we cannot reliably distinguish between patients were lost to care and those who were lost our data. Another consideration is that receiving MOUD at discharge was not necessarily associated with a lower rate of acute care return. This fragmentation likely reflects the absence of a single accountable pathway spanning acute and outpatient settings where each transition depends on a discrete hand-off (prescribing, scheduling, patient follow-through, clinic capacity).

## Limitations

These findings should be evaluated while acknowledging some specific limitations of our methods. First, as this data was collected from an electronic medical record, completeness and quality of the medical records cannot be verified. That being said, we are using this data as a surrogate for screening, treatment and referral or follow-up process opportunities. We believe that we are missing diagnosis codes for many patients who have not yet received a diagnosis code of opioid use disorder (OUD population, n=5,390), far more so than those who have an erroneous or no longer appropriate diagnosis code, and as such believe these to be underestimates of intervention opportunities. We believe that the absence of a diagnose code during an acute visit among those with known OUD (n=5390) often does not represent the absence of relevant active OUD, but rather a missed opportunity in acknowledgement and treatment of a relevant comorbid disorder. We had to integrate both medication reconciliation and prescription data as both are not always updated and might provide erroneous or not up to date data. While encounters with an OUD diagnosis code are not always for an OUD, their inclusion in this data supports the complexity of medical issues and the healthcare utilization burden of this population. In reverse, there are many encounters, especially in the Emergency Department, where secondary and tertiary codes are not listed and as such, opioid use disorder, its screening and or management might not have been included. That said, based on other prescription datasets we know our ED based buprenorphine/buprenorphine-naloxone ordering practices and know they are rare, and as such, we anticipate this limitation only underestimates the opportunities for improvement in treatment services. Overall, we did not see disparities in our prescribing practices. This data should be taken with some caution, because in general, we have just such low rates of prescriptions across all subgroups. The population with OUD in general is a population which is at risk for bias and inequitable care, separate from health inequities related to other socio-demographics; one inequity might be masking the other.

## Data Availability

All data produced in the present study are available upon reasonable request to the authors

## Ethics Review and Approval

This study was conducted as a Quality Improvement initiative and received approval from the Duke Institutional Review Board (Pro 00118790). The IRB issued the determination that the study is exempt from a full IRD review as it does not constitute human subject research. As this was a cohort study with de-identified data extracted from the electronic medical record, a waiver of informed consent was granted. All measures were taken throughout to ensure confidentiality and data security.

## Appendix Tables

**Table 1:** Patients with multiple index encounters for with an acute OUD encounter.

| Patients with a DUH/DRH acute OUD encounter<br>(Population 1)<br>N = 2,267 |  |
| --- | --- |
| Number of DUH/DRH acute OUD encounters |  |
| 1 | 1,580 (69.7%) |
| 2 | 361 (15.9%) |
| 3 | 117 (5.2%) |
| 4 | 86 (3.8%) |
| 5-10 | 109 (4.8%) |
| 11-15 | 12 (0.5%) |
| >15 | 2 (0.1%) |

**Table 2:** Demographic and clinical characteristics for patients with an inpatient or ED visit.

|  | Overall<br>N = 2,267 | Patients with at<br>least one inpatient<br>acute OUD<br>encounter<br>N = 1,837 | Patients with ED<br>acute OUD<br>encounters only<br>N = | ASMD |
| --- | --- | --- | --- | --- |
| **Demographics** |  |  |  |  |
| Age |  |  |  | 0.51 |
| Mean (SD) | 45.4 (14.4) | 46.7 (14.5) | 39.8 (12.9) |  |
| Median (Q1, Q3) | 43.3 (34.0, 56.2) | 45.0 (35.1, 57.6) | 37.4 (30.2, 47.7) |  |
| Min, Max | 18.0, 93.1 | 18.0, 93.1 | 18.0, 89.8 |  |
| Age - categorical |  |  |  | 0.30 |
| <20 | 31 (1.4%) | 19 (1.0%) | 12 (2.8%) |  |
| 20-29 | 285 (12.6%) | 195 (10.6%) | 90 (20.9%) |  |
| 30-39 | 627 (27.7%) | 480 (26.1%) | 147 (34.2%) |  |
| 40-49 | 497 (21.9%) | 407 (22.2%) | 90 (20.9%) |  |
| 50-59 | 413 (18.2%) | 355 (19.3%) | 58 (13.5%) |  |
| 60-69 | 283 (12.5%) | 260 (14.2%) | 23 (5.3%) |  |
| 70 and above | 131 (5.8%) | 121 (6.6%) | 10 (2.3%) |  |
| Sex |  |  |  | 0.17 |
| Female | 1,005 (44.3%) | 843 (45.9%) | 162 (37.7%) |  |
| Male | 1,262 (55.7%) | 994 (54.1%) | 268 (62.3%) |  |
| Race |  |  |  | 0.21 |
| American Indian or Alaska Native | 22 (1.0%) | 19 (1.0%) | 3 (0.7%) |  |
| Asian | 10 (0.4%) | 10 (0.5%) | 0 (0.0%) |  |
| Black | 792 (34.9%) | 615 (33.5%) | 177 (41.2%) |  |
| Native Hawaiian or Other Pacific<br>Islander | 1 (0.0%) | 1 (0.1%) | 0 (0.0%) |  |
| White | 1,347 (59.4%) | 1,127 (61.4%) | 220 (51.2%) |  |
| Other | 40 (1.8%) | 24 (1.3%) | 16 (3.7%) |  |
| Unknown | 55 (2.4%) | 41 (2.2%) | 14 (3.3%) |  |
| Hispanic ethnicity |  |  |  | 0.13 |
| Yes | 74 (3.3%) | 51 (2.8%) | 23 (5.3%) |  |
| No | 2,126 (93.8%) | 1,732 (94.3%) | 394 (91.6%) |  |
| Unknown | 67 (3.0%) | 54 (2.9%) | 13 (3.0%) |  |
| Race/ethnicity |  |  |  | 0.19 |
| Hispanic | 74 (3.3%) | 51 (2.8%) | 23 (5.3%) |  |
| Non-Hispanic Asian | 10 (0.4%) | 10 (0.5%) | 0 (0.0%) |  |
| Non-Hispanic Black | 773 (34.1%) | 603 (32.8%) | 170 (39.5%) |  |
| Non-Hispanic White | 1,286 (56.7%) | 1,075 (58.5%) | 211 (49.1%) |  |
| Non-Hispanic Other | 36 (1.6%) | 27 (1.5%) | 9 (2.1%) |  |
| Unknown | 88 (3.9%) | 71 (3.9%) | 17 (4.0%) |  |
| Patient preferred language |  |  |  | 0.10 |
| English | 2,253 (99.4%) | 1,829 (99.6%) | 424 (98.6%) |  |
| Spanish | 11 (0.5%) | 6 (0.3%) | 5 (1.2%) |  |
| Other | 3 (0.1%) | 2 (0.1%) | 1 (0.2%) |  |
| Insurance |  |  |  | 0.53 |
| Medicare | 560 (24.7%) | 502 (27.3%) | 58 (13.5%) |  |
| Medicaid | 1,025 (45.2%) | 841 (45.8%) | 184 (42.8%) |  |
| Other public | 16 (0.7%) | 14 (0.8%) | 2 (0.5%) |  |
| Private | 328 (14.5%) | 266 (14.5%) | 62 (14.4%) |  |
| Self-pay | 247 (10.9%) | 133 (7.2%) | 114 (26.5%) |  |
| Other | 91 (4.0%) | 81 (4.4%) | 10 (2.3%) |  |
| Missing | 0 (0.0%) | 0 (0.0%) | 0 (0.0%) |  |
| County of residence |  |  |  | 0.32 |
| Durham | 1,067 (47.1%) | 817 (44.5%) | 250 (58.1%) |  |
| Adjacent to Durham (Granville,<br>Wake, Chatham, Orange, Person) | 568 (25.1%) | 458 (24.9%) | 110 (25.6%) |  |
| Other NC county | 504 (22.2%) | 451 (24.6%) | 53 (12.3%) |  |
| Out of state | 79 (3.5%) | 66 (3.6%) | 13 (3.0%) |  |
| Missing | 49 (2.2%) | 45 (2.4%) | 4 (0.9%) |  |
| ADI |  |  |  | 0.15 |
| 1-5 | 1,091 (48.1%) | 859 (46.8%) | 232 (54.0%) |  |
| 6-10 | 963 (42.5%) | 806 (43.9%) | 157 (36.5%) |  |
| Missing | 213 (9.4%) | 172 (9.4%) | 41 (9.5%) |  |
| **Diagnoses** |  |  |  |  |
| HIV | 50 (2.2%) | 47 (2.6%) | 3 (0.7%) | 0.15 |
| Hep C | 421 (18.6%) | 399 (21.7%) | 22 (5.1%) | 0.50 |
| Psychiatric condition | 1,425 (62.9%) | 1,228 (66.8%) | 197 (45.8%) | 0.43 |
| Pain | 1,771 (78.1%) | 1,499 (81.6%) | 272 (63.3%) | 0.42 |
| Complications associated with<br>injected drug use* | 1,364 (60.2%) | 1,215 (66.1%) | 149 (34.7%) | 0.66 |
| Opioid overdose | 304 (13.4%) | 251 (13.7%) | 53 (12.3%) | 0.04 |
| Liver disease | 437 (19.3%) | 422 (23.0%) | 15 (3.5%) | 0.60 |
| Renal dysfunction | 829 (36.6%) | 790 (43.0%) | 39 (9.1%) | 0.84 |
| Cancer | 205 (9.0%) | 194 (10.6%) | 11 (2.6%) | 0.33 |
| Comorbidity score |  |  |  | 0.87 |

|  | **Overall**<br>N = 2,267 | **Patients with at least one inpatient acute OUD encounter**<br>N = 1,837 | **Patients with ED acute OUD encounters only**<br>N = | **ASMD** |
| --- | --- | --- | --- | --- |
| 0-2 | 888 (39.2%) | 580 (31.6%) | 308 (71.6%) |  |
| 3-4 | 962 (42.4%) | 853 (46.4%) | 109 (25.3%) |  |
| 5-8 | 417 (18.4%) | 404 (22.0%) | 13 (3.0%) |  |

**Table 3:** Rates of bup or bup-nalox prescription at discharge across comparator groups.

|  | **Bup or bup-nalox prescription at discharge**<br>N = 1,348 | **No bup or bup-nalox prescription at discharge**<br>N = 2,545 | **Acute DUH/DRH OUD encounters (Population 1 index encounters)**<br>N = 3,893 |
| --- | --- | --- | --- |
| Age |  |  |  |
| <30 | 147 (26.5%) | 408 (73.5%) | 555 (14.3%) |
| 30 and above | 1,201 (36.0%) | 2,137 (64.0%) | 3,338 (85.7%) |
| Sex |  |  |  |
| Female | 571 (33.2%) | 1,151 (66.8%) | 1,722 (44.2%) |
| Male | 777 (35.8%) | 1,394 (64.2%) | 2,171 (55.8%) |
| Race/ethnicity |  |  |  |
| Hispanic | 32 (30.2%) | 74 (69.8%) | 106 (2.7%) |
| Non-Hispanic Black | 473 (32.5%) | 983 (67.5%) | 1,456 (37.4%) |
| Non-Hispanic White | 768 (35.8%) | 1,376 (64.2%) | 2,144 (55.1%) |
| Other/Unknown | 75 (40.1%) | 112 (59.9%) | 187 (4.8%) |
| Insurance |  |  |  |
| Public | 997 (33.8%) | 1,953 (66.2%) | 2,950 (75.8%) |
| Private | 185 (40.1%) | 276 (59.9%) | 461 (11.8%) |
| Self-pay | 114 (33.3%) | 228 (66.7%) | 342 (8.8%) |
| Other | 52 (37.1%) | 88 (62.9%) | 140 (3.6%) |
| ADI |  |  |  |
| 1-5 | 502 (33.4%) | 1,002 (66.6%) | 1,504 (38.6%) |
| 6-10 | 507 (38.4%) | 813 (61.6%) | 1,320 (33.9%) |
| Missing | 339 (31.7%) | 730 (68.3%) | 1,069 (27.5%) |
| Encounter type |  |  |  |
| Inpatient | 1,089 (37.6%) | 1,807 (62.4%) | 2,896 (74.4%) |
| ED | 259 (26.0%) | 738 (74.0%) | 997 (25.6%) |
| Comorbidity score |  |  |  |
| 0-2 | 387 (33.8%) | 757 (66.2%) | 1,144 (29.4%) |
| 3-4 | 557 (33.0%) | 1,130 (67.0%) | 1,687 (43.3%) |
| 5-9 | 404 (38.0%) | 658 (62.0%) | 1,062 (27.3%) |

**Table 4:** Rates of acute care return visit within 30 days across comparator groups.

|  | Acute care return visit<br>within 30 days<br>N = 1,307 | No acute care return<br>visit within 30 days<br>N = 2,586 | Acute DUH/DRH OUD<br>encounters<br>(Population 1 index<br>encounters)<br>N = 3,893 |
| --- | --- | --- | --- |
| Age |  |  |  |
| <30 | 196 (35.3%) | 359 (64.7%) | 555 (14.3%) |
| 30 and above | 1,111 (33.3%) | 2,227 (66.7%) | 3,338 (85.7%) |
| Sex |  |  |  |
| Female | 580 (33.7%) | 1,142 (66.3%) | 1,722 (44.2%) |
| Male | 727 (33.5%) | 1,444 (66.5%) | 2,171 (55.8%) |
| Race/ethnicity |  |  |  |
| Hispanic | 31 (29.2%) | 75 (70.8%) | 106 (2.7%) |
| Non-Hispanic Black | 547 (37.6%) | 909 (62.4%) | 1,456 (37.4%) |
| Non-Hispanic White | 686 (32.0%) | 1,458 (68.0%) | 2,144 (55.1%) |
| Other/Unknown | 43 (23.0%) | 144 (77.0%) | 187 (4.8%) |
| Insurance |  |  |  |
| Public | 1,062 (36.0%) | 1,888 (64.0%) | 2,950 (75.8%) |
| Private | 120 (26.0%) | 341 (74.0%) | 461 (11.8%) |
| Self-pay | 86 (25.1%) | 256 (74.9%) | 342 (8.8%) |
| Other | 39 (27.9%) | 101 (72.1%) | 140 (3.6%) |
| ADI |  |  |  |
| 1-5 | 569 (37.8%) | 935 (62.2%) | 1,504 (38.6%) |
| 6-10 | 446 (33.8%) | 874 (66.2%) | 1,320 (33.9%) |
| Missing | 292 (27.3%) | 777 (72.7%) | 1,069 (27.5%) |
| Encounter type |  |  |  |
| Inpatient | 896 (30.9%) | 2,000 (69.1%) | 2,896 (74.4%) |
| ED | 411 (41.2%) | 586 (58.8%) | 997 (25.6%) |
| Comorbidity score |  |  |  |
| 0-2 | 238 (20.8%) | 906 (79.2%) | 1,144 (29.4%) |
| 3-4 | 595 (35.3%) | 1,092 (64.7%) | 1,687 (43.3%) |
| 5-8 | 474 (44.6%) | 588 (55.4%) | 1,062 (27.3%) |
| Bup or bup-nalox |  |  |  |
| Yes | 446 (33.1%) | 902 (66.9%) | 1,348 (34.6%) |
| No | 861 (33.8%) | 1,684 (66.2%) | 2,545 (65.4%) |

## Reference List

1. Harris MTH, Weinstein ZM, Walley AY. Medications for Opioid Use Disorder, Opioid Withdrawal, and Opioid Overdose. JAMA. 2026;335(11):986. doi:10.1001/jama.2025.26348

2. U.S. Overdose Deaths Decrease Almost 27% in 2024. 5/14/2025, 2025. https://www.cdc.gov/nchs/pressroom/releases/20250514.html?utm_source=chatgpt.com

3. Singh JA, Cleveland JD. National U.S. time-trends in opioid use disorder hospitalizations and associated healthcare utilization and mortality. PLoS One. 2020;15(2):e0229174. doi:10.1371/journal.pone.0229174

4. National Center for Health Statistics. U.S. overdose deaths decrease almost 27% in 2024. Centers for Disease Control and Prevention. Published May 14, 2025. Accessed September 23, 2026. https://www.cdc.gov/nchs/pressroom/releases/20250514.html

5. National Center for Health Statistics. U.S. overdose deaths decrease for third consecutive year in 2025. Centers for Disease Control and Prevention. Published May 13, 2026. Accessed September 23, 2026. https://www.cdc.gov/nchs/pressroom/releases/20260513.html

6. Murphy SM. The cost of opioid use disorder and the value of aversion. Drug Alcohol Depend. 2020 Dec 1;217:108382. doi: 10.1016/j.drugalcdep.2020.108382. Epub 2020 Oct 26. PMID: 33183909; PMCID: PMC7737485.

7. Qeadan F, Nicolson A, Tingey B, Moffett ML, Azagba S. Healthcare utilization trends among patients with opioid use disorder in U.S. Hospitals: an analysis of length of stay, total charges, and costs, 2005-2020. BMC Health Serv Res. 2025 Jul 4;25(1):927. doi: 10.1186/s12913-025-13095-9. PMID: 40615891; PMCID: PMC12226882.

8. Rosen AD, Takada S, Juillard C, Fernandez Montero YL, Richards AM, Ngekeng S, et al. Unpacking the link between substance use disorders and 30-day unplanned readmission. Addiction. 2025; 120(12): 2538–2546. 10.1111/add.70136

9. Langabeer JR, Stotts AL, Bobrow BJ, et al. Prevalence and charges of opioid-related visits to U.S. emergency departments. Drug Alcohol Depend. Apr 1 2021;221:108568. doi:10.1016/j.drugalcdep.2021.108568

10. Rosenthal ES, Brokus C, Sun J, Carpenter JE, Catalanotti J, Eaton EF, Steck AR, Kuo I, Burkholder GA, Akselrod H, McGonigle K, Moran T, Mai W, Notis M, Del Rio C, Greenberg A, Saag MS, Kottilil S, Masur H, Kattakuzhy S. Undertreatment of opioid use disorder in patients hospitalized with injection drug use-associated infections. AIDS. 2023 Oct 1;37(12):1799–1809. doi: 10.1097/QAD.0000000000003629. Epub 2023 Jun 20. PMID: 37352497; PMCID: PMC10481931.

11. O’Rourke BP, Hogan TH, Teater J, Fried M, Williams M, Miller A, Clark AD, Huynh P, Kauffman E, Hefner JL. Initiation of medication for opioid use disorder across a health system: A retrospective analysis of patient characteristics and inpatient outcomes. Drug Alcohol Depend Rep. 2022 Nov 12;5:100114. doi: 10.1016/j.dadr.2022.100114. PMID: 36844164; PMCID: PMC9948916.

12. Fiellin DA, O’Connor PG, Chawarski M, Pakes JP, Pantalon MV, Schottenfeld RS. Methadone maintenance in primary care: A randomized controlled trial. JAMA. 2001;286(14):1724–1731

13. Larochelle MR, Bernson D, Land T, et al. Medication for Opioid Use Disorder After Nonfatal Opioid Overdose and Association With Mortality. Annals of Internal Medicine. 2018;169(3):137–145. doi:10.7326/M17-3107

14. Dowell D, Brown S, Gyawali S, et al. Treatment for Opioid Use Disorder: Population Estimates — United States, 2022. MMWR Morbidity and Mortality Weekly Report. 2024;73(25):567–574. doi:10.15585/mmwr.mm7325a1

15. Barnett Michael L, Meara E, Lewinson T, et al. Racial Inequality in Receipt of Medications for Opioid Use Disorder. New England Journal of Medicine. 2023/05/10 2023;388(19):1779–1789. doi:10.1056/NEJMsa2212412

16. Chan B, Ezekiel-Herrera D, Bailey SR, et al. Race, Ethnicity, and Language Disparities in Alcohol and Drug Screening and Medication Treatment. JAMA Network Open. 2026;9(5):e2612319. doi:10.1001/jamanetworkopen.2026.12319

17. Dunphy CC, Zhang K, Xu L, Guy GP, Jr. Racial‒Ethnic Disparities of Buprenorphine and Vivitrol Receipt in Medicaid. American Journal of Preventive Medicine. 2022;63(5):717–725. doi:10.1016/j.amepre.2022.05.006

18. Highlights for the 2022 National Survey on Drug Use and Health. Substance Abuse and Mental Health Services Administration (SAMHSA). https://www.samhsa.gov/data/sites/default/files/reports/rpt42731/2022-nsduh-main-highlights.pdf

19. Busch SH, Fiellin DA, Chawarski MC, et al. Cost-effectiveness of emergency department-initiated treatment for opioid dependence. Addiction. 2017;112(11):2002–2010. 10.1111/add.13900

20. Barocas JA, Savinkina A, Adams J, et al. Clinical impact, costs, and cost-effectiveness of hospital-based strategies for addressing the US opioid epidemic: a modelling study. The Lancet Public Health. 2022;7(1):e56–e64. doi:10.1016/S2468-2667(21)00248-6

21. D’Onofrio G, O’Connor PG, Pantalon MV, et al. Emergency Department–Initiated Buprenorphine/Naloxone Treatment for Opioid Dependence. JAMA. 2015;313(16):1636. doi:10.1001/jama.2015.3474

22. Englander H, Dobbertin K, Lind BK, et al. Inpatient Addiction Medicine Consultation and Post-Hospital Substance Use Disorder Treatment Engagement: a Propensity-Matched Analysis. Journal of General Internal Medicine. 2019/12/01 2019;34(12):2796–2803. doi:10.1007/s11606-019-05251-9

23. Regan S, Howard S, Powell E, et al. Emergency Department-initiated Buprenorphine and Referral to Follow-up Addiction Care: A Program Description. Journal of Addiction Medicine. 2021/06/17 2022;16(2):216–222. doi:10.1097/ADM.0000000000000875

24. Wakeman SE, McGovern S, Kehoe L, et al. Predictors of engagement and retention in care at a low-threshold substance use disorder bridge clinic. J Subst Abuse Treat. Oct 2022;141:108848. doi:10.1016/j.jsat.2022.108848

25. Wakeman SE, Metlay JP, Chang Y, Herman GE, Rigotti NA. Inpatient Addiction Consultation for Hospitalized Patients Increases Post-Discharge Abstinence and Reduces Addiction Severity. Journal of General Internal Medicine. 2017/08/01 2017;32(8):909–916. doi:10.1007/s11606-017-4077-z

26. Weiner SG, Little K, Yoo J, et al. Opioid Overdose After Medication for Opioid Use Disorder Initiation Following Hospitalization or ED Visit. JAMA Network Open. 2024;7(7):e2423954. doi:10.1001/jamanetworkopen.2024.23954

27. Kind AJH, Buckingham W. Making Neighborhood Disadvantage Metrics Accessible: The Neighborhood Atlas. New England Journal of Medicine, 2018. 378: 2456–2458. DOI: 10.1056/NEJMp1802313. PMCID: PMC6051533.

28. University of Wisconsin School of Medicine Public Health. 2015 Area Deprivation Index v2.0. Downloaded from https://www.neighborhoodatlas.medicine.wisc.edu/ May 23, 2019.

29. Rostam-Abadi Y, Wang S, King C, Kalyanaraman Marcello R, Van Wye G, Tuazon E, Kennedy J, Cooke C, Mazumdar M, Tarpey T, Billings J, Appleton N, Fernando J, Fawole A, Siddiqui S, Barron C, Schatz D, McNeely J. Addiction Consult Services, Mortality, and Acute Care Utilization in Inpatients With Opioid Use Disorder: A Secondary Analysis of a Cluster Randomized Clinical Trial. JAMA Netw Open. 2025 Aug 1;8(8):e2525222. doi: 10.1001/jamanetworkopen.2025.25222. PMID: 40768148; PMCID: PMC12329607.

30. Skains RM, Reynolds L, Carlisle N, Heath S, Covington W, Hornbuckle K, Walter L. Impact of Emergency Department-Initiated Buprenorphine on Repeat Emergency Department Utilization. West J Emerg Med. 2023 Nov;24(6):1010–1017. doi: 10.5811/westjem.60511. PMID: 38165181; PMCID: PMC10754187.

31. Armour R, Nielsen S, Buxton JA, Bolster J, Han MX, Ross L. Initiation of buprenorphine in the emergency department or emergency out-of-hospital setting: A mixed-methods systematic review. Am J Emerg Med. 2025 Feb;88:12–22. doi: 10.1016/j.ajem.2024.11.031. Epub 2024 Nov 17. PMID: 39577213.

32. Clifton D, Ivey N, Platt A, Hong C, Setji N. Outcomes of a Hospitalist-Led Consult Service for Patients with Opioid Use Disorder: A Propensity Score Weighted Study. J Gen Intern Med. 2026 Jan;41(2):287–295. doi: 10.1007/s11606-025-09820-z. Epub 2025 Nov 6. PMID: 41198992; PMCID: PMC12894605.

33. Hughes T, Nasser N, Mitra A. Overview of best practices for buprenorphine initiation in the emergency department. Int J Emerg Med. 2024 Feb 19;17(1):23. doi: 10.1186/s12245-024-00593-6. PMID: 38373992; PMCID: PMC10877824.

34. Seliski N, Madsen T, Eley S, Colosimo J, Engar T, Gordon A, Barnett C, Humiston G, Morsillo T, Stolebarger L, Smid MC, Cochran G. Implementation of a rural emergency department-initiated buprenorphine program in the mountain west: a study protocol. Addict Sci Clin Pract. 2024 Sep 3;19(1):63. doi: 10.1186/s13722-024-00496-0. PMID: 39228007; PMCID: PMC11369999.

35. Wilder ME, Richardson LD, Hoffman RS, Winkel G, Manini AF. Racial disparities in the treatment of acute overdose in the emergency department. Clin Toxicol (Phila). 2018 Dec;56(12):1173–1178. doi: 10.1080/15563650.2018.1478425. Epub 2018 Jun 12. PMID: 29893609; PMCID: PMC6318059.

36. Scott KW, Scott JW, Sabbatini AK, Chen C, Liu A, Dieleman JL, Duber HC. Assessing Catastrophic Health Expenditures Among Uninsured People Who Seek Care in US Hospital-Based Emergency Departments. JAMA Health Forum. 2021 Dec 30;2(12):e214359. doi: 10.1001/jamahealthforum.2021.4359. PMID: 35977304; PMCID: PMC8796980.

37. Madden EF, Prevedel S, Light T, Sulzer SH. Intervention Stigma toward Medications for Opioid Use Disorder: A Systematic Review. Subst Use Misuse. 2021;56(14):2181–2201. doi: 10.1080/10826084.2021.1975749. Epub 2021 Sep 20. PMID: 34538213.

38. Cheetham A, Picco L, Barnett A, Lubman DI, Nielsen S. The Impact of Stigma on People with Opioid Use Disorder, Opioid Treatment, and Policy. Subst Abuse Rehabil. 2022 Jan 25;13:1–12. doi: 10.2147/SAR.S304566. PMID: 35115860; PMCID: PMC8800858.

39. Quadri OH, Skogseth E, Brant K, Jones AA. The role of micro-, meso-, and macro-level stigma on the uptake of medications for opioid use disorder (MOUD) among women in the criminal legal system. BMC Womens Health. 2025 Jul 25;25(1):370. doi: 10.1186/s12905-025-03863-4. PMID: 40713527; PMCID: PMC12291414.

